# Outcomes of the Advanced Visualization In Corneal Surgery Evaluation (ADVISE) trial; a non-inferiority randomized control trial to evaluate the use of intraoperative OCT during Descemet membrane endothelial keratoplasty

**DOI:** 10.1101/2022.01.18.22269460

**Authors:** Marc B Muijzer, H. Delbeke, M.M. Dickman, R.M.M.A. Nuijts, H.J. Noordmans, S.M. Imhof, Robert P L Wisse

## Abstract

**Purpose:** To evaluate if an intraoperative OCT (iOCT) optimized surgical protocol without prolonged overpressure is non-inferior to a standard protocol during Descemet membrane endothelial keratoplasty (DMEK).

**Design:** A multicenter international prospective non-inferiority randomized control trial

**Subjects:** Sixty-five pseudophakic eyes of 65 patients with corneal endothelial dysfunction resulting from Fuchs endothelial corneal dystrophy were enrolled in 3 corneal centers in The Netherlands and Belgium.

**Methods:** The study was powered to include 63 patients scheduled for routine DMEK. Subjects were randomized to the control arm (n=33) without iOCT-use and raising the intraocular pressure above normal physiological limits for 8 minutes (i.e., overpressure) or the intervention arm (n=32) with OCT-guidance to assess graft orientation and adherence while refraining from prolonged raising the intraocular pressure. The RD and 95% confidence intervals (95% CI) were calculated from a logistic regression model using 1,000 bootstrap samples. Secondary outcomes included the incidence of graft detachment, surgeon-reported iOCT-aided surgical decision making, surgical time, endothelial cell density (ECD), and corrected distance visual acuity (CDVA).

**Main Outcome Measures:** The primary outcome was the incidence of postoperative surgery-related adverse events, defined as rebubbling, graft failure, and iatrogenic acute glaucoma. The non-inferiority margin was set at a risk difference (RD) of 10%.

**Results:** In the control group, 13 adverse events were recorded in 10 subjects compared to 13 adverse events in 12 subjects in the intervention group. The mean unadjusted RD measured 0.38% (95%CI: - 9.64–10.64) and the RD adjusted for study site measured -0.32% (95%CI: -10.29–9.84). No significant differences in ECD and CDVA were found between the two groups 3 and 6 months postoperatively. Surgeons reported that iOCT aided surgical decision-making in 40% of cases. Surgical- and graft unfolding time were, respectively, 13% and 27% shorter in the iOCT-group.

**Conclusions:** iOCT-guided DMEK surgery with refraining from prolonged over-pressuring was non-inferior compared to conventional treatment. Surgery times were reduced considerably, and surgeons reported the iOCT aided surgical decision-making in 40% of cases. Refraining from prolonged overpressure did not affect postoperative ECD or CDVA.

## Introduction

Intraoperative-OCT (iOCT) provides surgeons with real-time feedback to assess surgical events, anatomical changes, and surgical manipulations, otherwise not possible using the *en face* view of the surgical microscope.^1^ Numerous studies described the value of this emergent technology in ophthalmic surgery.^1,2^ In particular, the iOCT aids clinical-decision making, enables surgeons to in-vivo study their surgical practice patterns, and achieving a greater understanding of pathophysiology and surgical tissue alterations. Nevertheless, most previous studies were observational, had small sample sizes and lacked a control group making it difficult to quantify the putative benefits of iOCT.

One promising surgery to reap the benefits of iOCT is Descemet membrane endothelial keratoplasty (DMEK).^3–6^ DMEK is a recent iteration of endothelial keratoplasty and reported advantages include faster visual recovery, superior visual acuity, and reduced rates of endothelial rejection compared to Descemet stripping endothelial keratoplasty.^7–9^ Despite these advantages the rate of postoperative adverse events (e.g., graft detachment requiring rebubbling) for DMEK is relatively high with a reported prevalence of ranging between 2% and 82% for rebubbling and 3% and 11% for primary graft failure.^9–12^ These adverse events necessitate secondary surgical interventions and are associated with a lower graft viability and survival.^10^

The causes of graft detachment and primary graft failure are considered multifactorial and can be divided into donor, patient, and surgical factors.^10,13^ The primary focus of current research considers modifications of surgical techniques to prevent these complications.^16–18^ Among other factors, graft adherence issues due to fluids between the donor and recipient, insufficient anterior chamber (AC) tamponade pressure, and graft trauma have been proposed to cause postoperative complications. Several techniques have been described to promote graft adherence, such as corneal swiping and prolonged intraoperative over-pressurising of the eye.^15,17^ There is no consensus about the best approach and prolonged overpressurizing the globe has been widely discussed.^16,18–21^ It has been theorized that prolonged overpressure may push residual interface fluid into the stroma and improve graft adherence resulting in lower rebubbling rates.^15,16,21^ Other research shows a limited effect on DMEK graft adherence.^19,20^ On the other hand, overpressure may lead to potential adverse side effects, including endothelial cell loss^16^, exacerbation of glaucoma^22^, and a compromised retinal perfusion.^23^

iOCT enables surgeons to directly assess graft adherence, the need for additional surgical manoeuvres, and facilitates DMEK orientation without the need for external markings that may damage the graft and increase the risk of complications.^5,6^ In the PIONEER and DISCOVER study Ehlers et al. reported that iOCT aided and altered clinical decision making in, respectively, 48 and 43% of corneal surgeries.^24,25^ The iOCT provided valuable feedback in evaluating graft-host apposition, graft positioning, and verifying graft orientation in DMEK. In addition, several studies show promising evidence iOCT enables faster positioning of the graft with fewer manipulations.^3–5^

These insights led to the conceptualization of an iOCT-optimized DMEK surgical protocol by our group, consisting of iOCT-guidance during unfolding and refraining from prolonged over-pressuring of the globe. In a pilot study, the incidence of postoperative adverse events was lower and operation time was shorter using this protocol.^6^ Notwithstanding, in this pilot protocol changes were gradually introduced and a control without iOCT guidance was missing. The promising results warranted follow-up in a head-to-head comparison with a conventional surgical protocol. In this study we investigate whether iOCT-guidance can obviate the need for prolonged overpressure in DMEK surgery and can be considered non-inferior to a standard protocol in terms of postoperative adverse events. Here, we present the results of our prospective *Advanced Visualization In Corneal Surgery Evaluation* (ADVISE), a non-inferiority randomized clinical trial designed to answer these questions.

## Materials and methods

### Study protocol

All subjects provided written informed consent and were included in the prospective *Advanced Visualization In Corneal Surgery Evaluation* (ADVISE) trial, an international non-inferiority single-blinded RCT to investigate the utility of intraoperative optical coherence tomography (OCT) in DMEK surgery. Subjects underwent surgery between December 2018 and April 2021 in the University Medical Center Utrecht (n = 39), University Hospital Leuven (n = 14), or Maastricht University Medical Center (n= 14).

Inclusion criteria were pseudophakic adult patients with irreversible corneal endothelial dysfunction resulting from Fuchs endothelial corneal dystrophy eligible for DMEK surgery. Exclusion criteria were human-leukocyte antigen matched keratoplasty, any ocular comorbidity other than ocular surface disease, open angle glaucoma, and mild age-related macular degeneration. No combined phaco-emulsification procedures were performed and only one eye per subject was enrolled. Subjects were randomized to either the iOCT-group or control group using minimization randomization stratified for center using an embedded function of the Electronic Data Capture platform (Research Online, Julius Center, Utrecht, The Netherlands). Patients were blinded throughout the study period. The surgeons and researchers could not be blinded, as the surgeons performed the surgery and researchers were present during surgery to facilitate imaging.

All procedures were performed in accordance with the Declaration of Helsinki, local and national laws regarding research (i.e., the Act on Scientific Research Involving Humans), European directives with respect to privacy (General Data Protection Regulation 2016/679), and 2010 CONSORT standards for reporting RCT’s.^26^ The study was approved by the Ethics Review Boards in The Netherlands and Belgium (Medical Ethics Committee Utrecht file no. 18-487, Ethical committee Leuven file no. S61527) and registered at clinicaltrials.gov (number: NCT03763721) and CCMO.nl (number: NL64392.041.17).

### Study measurements

Each patient underwent an ophthalmic examination preoperatively and 1 day, 1 week, 1 month, 3 months, and 6 months after surgery. Here, we report the baseline, 3 months, 6 months, and all adverse events in detail. The ophthalmic examinations included a full slit-lamp examination, fundus examination, intraocular pressure (IOP) measurement, Scheimpflug tomography (Pentacam HR type 70900, Oculus GmbH, Wetzlar, Germany), anterior segment OCT (Utrecht and Leuven: Zeiss Cirrus 5000, Zeiss Meditec, Oberkochen, Germany; Maastricht: Casia SS-1000, Tomey, Nagoya, Japan), and posterior segment OCT (Utrecht and Leuven: Zeiss Cirrus 5000, Zeiss Meditec, Oberkochen, Germany; Maastricht: Spectralis, Heidelberg Engineering GmbH, Heidelberg, Germany), and an endothelial cell count (EM4000, Tomey, Nagoya, Japan; SP-3000; Topcon, Nagoya, Japan). An optometrist measured the manifest refraction and the corrected distance visual acuity (CDVA) using an Early Treatment Diabetic Retinopathy Study (ETDRS) letter chart at 4 meters.

### Surgical procedure

Donor grafts were allocated by the Dutch Transplant Foundation (Nederlandse Transplantatie Stichting, Leiden, the Netherlands). The grafts were organ cultured and provided pre-stripped by the ETB-Bislife (Beverwijk, the Netherlands), with a minimum endothelial cell density (ECD) of 2300 cells/mm2 and with a diameter of 8.5 mm. All surgical procedures were performed by experienced corneal surgeons (H.D., R.M.M.A.N, M.M.D., R.P.L.W.), following a largely standardized procedure. Prior to surgery, 27 subjects underwent a Nd:YAG laser iridotomy at 6 o’clock according to the preference of the surgeon. In the other 38 subjects, a surgical iridectomy was performed using a 27-gauge needle and Price hook at 6 o’clock following the Descemetorhexis. In all cases a 2.8 mm corneal incision was made, followed by a 9 mm Descemetorhexis under air in 51 subjects and a viscoelastic device in 14 subjects (Healon; Abbott Medical, Uppsala, Sweden). The graft was stained using trypan blue dye (Membrane blue n = 52, Vision Blue, n = 13, both from DORC, Zuidland, the Netherlands) and inserted into the anterior chamber using a glass injector (Geuder AG, Heidelberg, Germany, n= 52, DORC, Zuidland, the Netherlands, n = 13). No touch technique used to unfold and position the graft.^27^ In 33 surgeries the randomization dictated that iOCT was not available to the surgeons. Here, a full AC fill was performed, raising the IOP above normal physiological limits for 8 minutes using air (*overpressure)*. In the other 32 surgeries the graft was positioned as described above and iOCT was available for utilization at the surgeon’s discretion during unfolding and used to check for complete adherence of the graft without overpressurizing the eye. At the end of surgery, the air was replaced by 20% Sulphur Hexafluoride gas and the size of the gas bubble was reduced to cover the graft (i.e., same size as the graft). Next, a validation scan of proper apposition by iOCT was performed in both the control and intervention arm, as proposed by the ethical review board. Any irregularities were treated at the discretion of the surgeon. After surgery, patients remained strictly supine for two hours at the hospital and were instructed to remain in the same position for the following 24 hours. All surgeons reported on the quality of the iOCT image and whether the iOCT aided surgical decision-making, such as unfolding the graft and determining orientation of the graft. All surgical videos were qualitatively analyzed by two graders (M.B.M. and an independent grader) to record graft unfolding grade, graft geometry, centering of the graft, and surgical times. Graft unfolding grade was classified in 4 grades depending on the required manipulation and time to unfold/position the graft as earlier described by Maier et al.^28^ Grade I refers to a primarily correct oriented graft after insertion in the AC with a straightforward and direct unfolding and centering of the graft. Grade II is described as slightly more complicated using indirect methods to unfold and center the graft (duration < 5 minutes). Grade III is difficult unfolding and centering using indirect methods requiring repeated air injection with BSS exchange (duration > 5 minutes). Grade IV refers to grade III with direct manipulation of the graft using a cannula or forceps for unfolding and centering. *Outcome measures:*

The primary outcome was the incidence of postoperative adverse events, defined as graft detachments requiring surgical intervention (i.e., rebubbling), primary graft failures, or iatrogenic acute glaucoma. Rebubbling was performed at the discretion of the surgeon, though principally when the graft was >30% detached or the detachment involved the visual axis. Secondary outcomes consisted of surgeon reported iOCT-aided surgical decision-making, a qualitative analysis of surgical video’s, surgical time, postoperative ECD loss, and CDVA at follow-up. Directly after the surgery the surgeons was asked on whether the iOCT-aided surgical decision making and if applicable how the iOCT-aided surgical decision making. The surgical time was recorded, and the time of various surgical steps was determined after surgery by manual review of the surgical video. Postoperative ECD loss was determined by calculating the difference between the donor graft ECD and the post-operative specular light microscopy assessments. The ETDRS letter score of the CDVA was converted to logarithm of the minimum angle of resolution (logMAR) units by multiplying the number of letters read by -0.02 log units and adding 1.7 log units.^29^

All graft detachments, defined as any non-adherence of the graft noticeable on slit lamp examination and AS-OCT imaging at any time point within 3 months after surgery, were recorded. Using a cornea grid consisting of 25 cornea zones, the presence and size of the graft detachments were quantified.

### Sample size

Power calculation was based on the incidence of postoperative adverse events. The non-inferiority limit was set at 10% and was set as a clinically relevant risk difference (RD), based on clinical judgment and available data at the time of trial design. Thus, non-inferiority would be demonstrated if the upper boundary of the 95% confidence interval (CI) of the RD between both treatment arms is lower than 10%. Assuming an α of 0.05 (1-sided) and a power of 80%, and a non-inferiority limit of 10%, a sample size of at least 60 subjects would be required (30 per treatment arm). Considering a loss to follow up of 5%, the final computed sample size was 63 subjects. The power calculation did not provide for COVID-19 related loss to follow-up (n=4).

### Statistical analysis

The primary dependent variable consisted of the total counted adverse events developed by each patient and converted to a proportion for analysis. For the analysis of the primary outcome measure a crude and adjusted marginal risk difference (RD) between the two treatment arms was estimated from a logistic regression model using 1,000 bootstrap samples.^30^ The primary analysis was adjusted treatment site to correct for differences in number of inclusions, unacknowledged differences in practice patterns, and surgeon experience. *P*-values cannot be calculated from the described method and only can be estimated using the 95% CI. A stratified analysis was performed to calculate the unadjusted RD for graft detachment, rebubbling, primary graft failure, and iatrogenic acute glaucoma. For a stratified adjusted analysis, it appeared not possible to calculate reliable estimates. A secondary regression analysis was performed to estimate the effect of overpressure duration in minutes on the incidence of graft detachment and area of detachment.

Missing observation of the secondary outcomes; CDVA, central cornea thickness, ECD, retinal nerve fiber layer thickness, and IOP, were imputed using multiple imputation. Missing measurements of subjects that developed a graft failure were considered missing not at random and not imputed. The other missing observations were considered missing at random. The variables concerned and baseline variables concerned were used as predictors for imputing. The number of imputations was equal to the maximum percentage of missing data plus one. All secondary outcomes were analyzed using the student t-test for differences between treatment arms. Correction for multiple comparisons was performed using the Bonferroni correction. A 2-sided *P* value < 0.05 was considered statistically significant.

An intention-to-treat analysis was performed for all outcomes measures. All statistical analysis were performed using R statistical software version 4.0.3 (Comprehensive R Archive Network, Vienna, Austria). All statistical analysis were supervised by an independent statistician from the Julius Center for health sciences (https://www.juliusclinical.com/). Data are described as mean ±standard deviation (SD) for continuous variables and as individual counts and percentages for dichotomous and categorical variables.

## Results

A total of 66 eyes of 66 patients were randomized to either the conventional protocol (control group, n = 33) or the iOCT-optimized protocol (intervention group, n = 33). One subject discontinued the study after randomization before receiving surgery and was replaced by a new subject. In the control group 2 crossover cases were recorded, in which the iOCT was used to salvage the graft in a complicated procedure. In both cases 8 minutes of overpressure was applied at the end of surgery. All remaining patients in both treatment arms received the allocated treatment. Four serious adverse events were recorded over the course of the study, 3 subjects underwent re-transplantation for primary graft failure and one subject included in the study died of multi-organ failure unrelated to the study before randomization. The deceased subject was subsequently excluded without replacement. In total, 7 subjects were lost to follow-up; 3 subjects dropped out after re-transplantation and 4 subjects were lost to follow-up because of reduction in care delivery caused by the COVID-19 pandemic. For all subjects who underwent surgery (n=65) the primary outcome was obtained and included for analysis (**Fig 1**.). Baseline patient and donor characteristics are displayed in Table 1. Commensurate with the 2012 CONSORT guidelines, baseline characteristics were not tested for statistical differences.^26^ During the study 5 surgical complications were recorded: 2 cases with endothelial damage due to graft manipulation and 3 cases with an anterior chamber hemorrhage.

**Figure 1.**
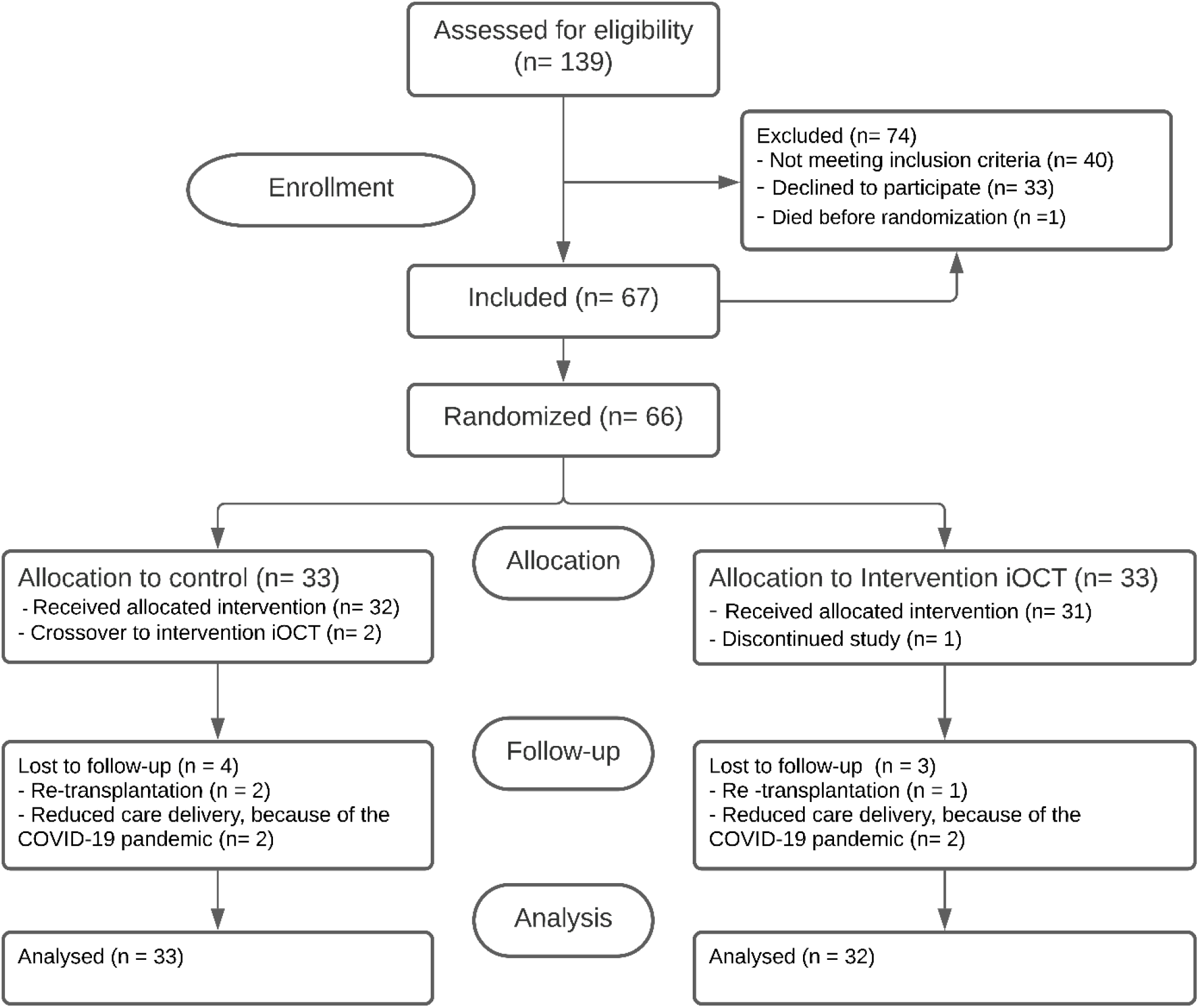
Consolidated standards of reporting trials flowchart.

**Table 1.** Baseline Patient and Donor Characteristics

|  | Conventional protocol (n=33) | iOCT-optimized protocol (n= 32) |
| --- | --- | --- |
| **patient** |  |  |
| Sex (female), n (%) | 17 (52) | 17 (53) |
| Age (years), mean (SD) | 72.4 (6.6) | 73.3 (6.4) |
| CDVA (logMAR), mean (SD) | 0.42 (0.25) | 0.41 (0.26) |
| Pachymetry (μm), mean (SD) | 625 (86) | 595 (62) |
| RFNL thickness (μm), mean (SD) | 89 (13) | 87 (13) |
| IOP (mmHg), mean (SD) | 12.9 (3.3) | 12.6 (3.0) |
| Corneal edema present, n (%) | 15 (45.5) | 13 (40.6) |
| Descemet folds present, n (%) | 2 (6.1) | 6 (18.8) |
| Bullae present, n (%) | 5 (15.2) | 4 (12.5) |
| Laser iridotomy, n (%) | 15 (45.5) | 12 (37.5) |
| **Donor** |  |  |
| Age (years), mean (SD) | 74.3 (5.0) | 73.3 (5.8) |
| ECD (cells/mm<sup>2</sup>), mean (SD) | 2706 (174) | 2719 (180) |

CDVA: corrected distance visual acuity; ECD: endothelial cell density; logMAR: logarithm of the minimum angle of resolution; IOP: intra-ocular pressure; SD: standard deviation; RFNL: retinal nerve fiber layer

### Incidence of postoperative adverse events and clinical outcomes

A total of 26 postoperative adverse events were recorded in 22 subjects (control group: 13 adverse events in 10 subjects, intervention group: 13 adverse events in 12 subjects). In the iOCT group 17 graft detachments were recorded resulting in 11 rebubbling procedures, compared to 16 detachments resulting in 6 rebubbling procedures in the control group. The area of detachment in cases requiring rebubbling measured 44% (SD ±25%) of the cornea surface in the iOCT group compared to 39% (SD ±10%) in the control group (P=0.655, 95%CI: 0.18 – 0.28). Three primary graft failures were recorded (control group n = 2, iOCT-group n = 1), all cases were preceded by a graft detachment and subsequent unsuccessful rebubbling of the graft. In the control group, 5 cases developed an iatrogenic pupillary block glaucoma in the first 24 hours after surgery compared to 1 case in the iOCT group. No statistically significant differences in the incidence of adverse events were found between the iOCT group and the control group (table 2).

**Table 2.**
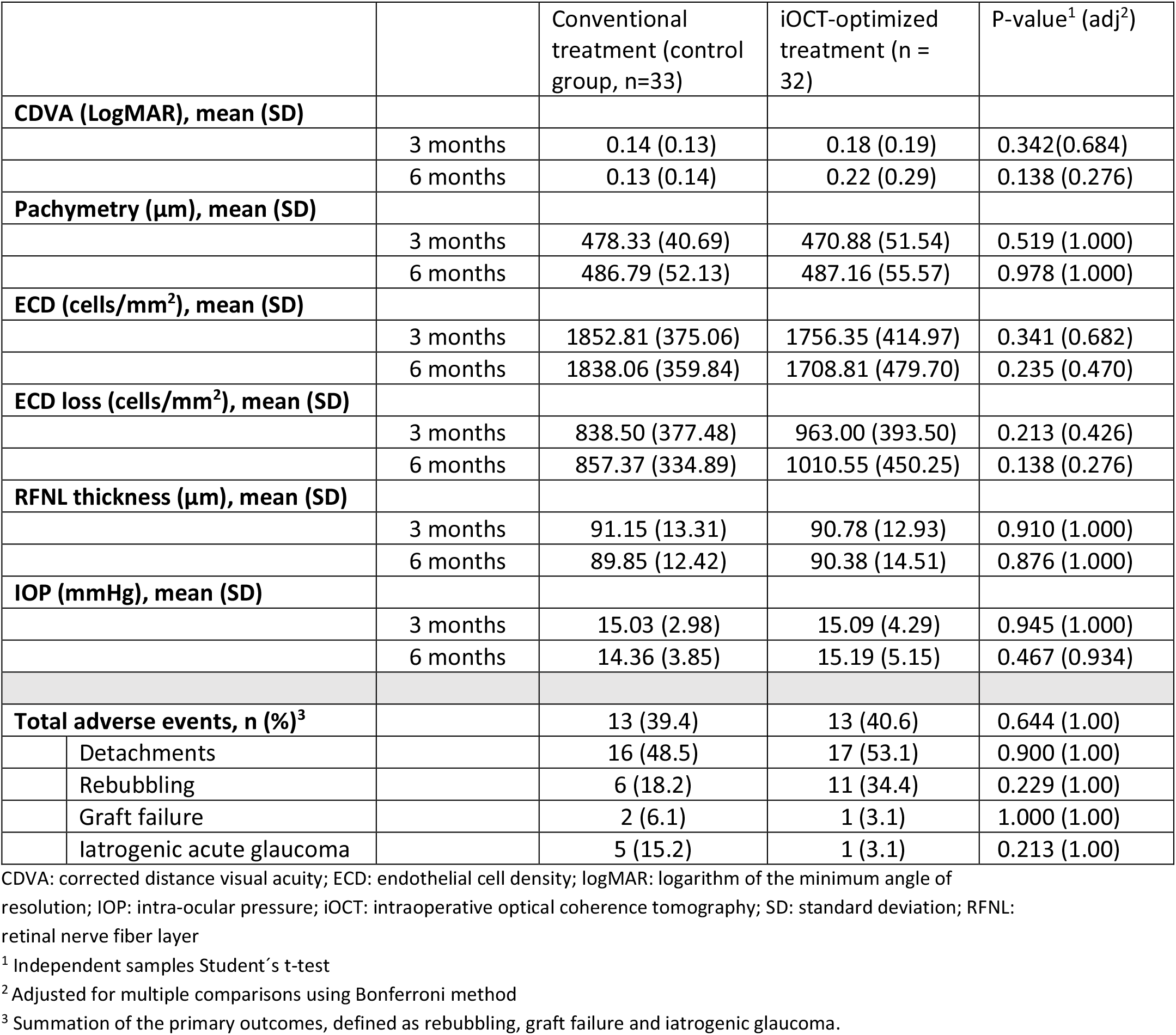
Primary and secondary outcomes after Conventional treatment and iOCT-optimized treatment

We explored unadjusted and adjusted estimates of the iOCT-optimized surgical protocol on the postoperative adverse event rate. The mean unadjusted risk difference (RD) measured 0.38% (95%CI: -9.64 – 10.64) and the RD adjusted for study site measured -0.32% (95%CI: -10.29 – 9.84), meaning in short that both protocols are comparable with regards to overall surgical safety measured as total postoperative adverse event rate. After controlling for a priori planned adjustment for study site, the iOCT-optimized protocol was found non-inferior to the conventional protocol (Figure 2). In addition, the independent effect of overpressure duration measured in minutes was not significantly associated with the incidence of detachment (β: 0.02, 95%CI: -0.10 – 0.15, *P*= 0.730) or area of detachment (β: - 0.012, 95%CI: -0.027 – -0.002, *P*= 0.121). The unadjusted and adjusted regression models of the primary outcome can be found in the supplementary data (Supplementary table 1).

**Figure 2.**
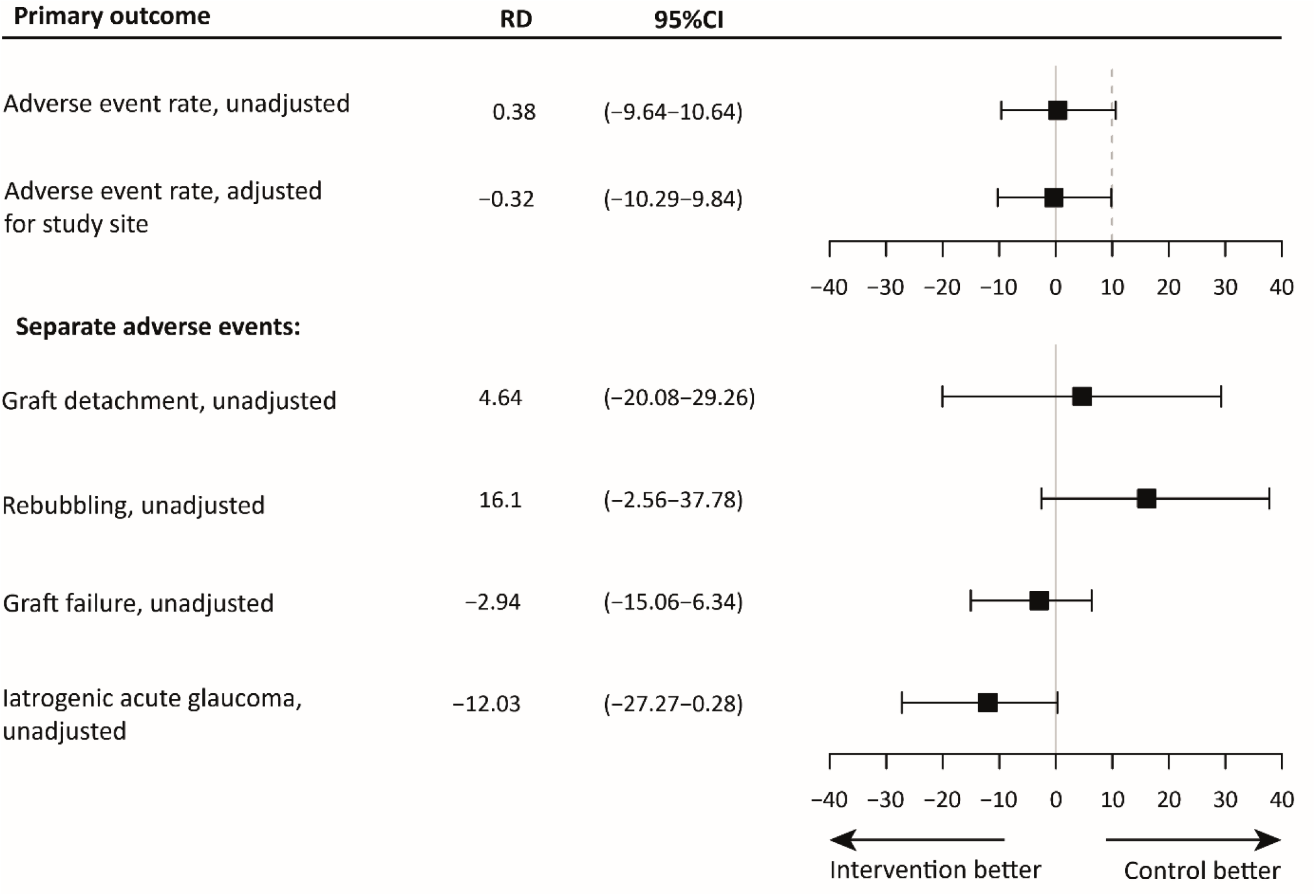
The mean risk difference (RD) and 95% confidence interval (CI) of the outcome measures, and the non-inferiority limit (dashed line). The top panel shows the unadjusted and adjusted estimates for the primary outcome measure. The bottom panel shows the unadjusted estimates for all separate postoperative events. For these outcomes, a non-inferiority margin is not shown.

When reporting individual adverse events, the results show varying results regarding the RDs. Consistent with the observed adverse events the risk of graft detachment (+4.6%) and rebubbling (+16.1%) are increased compared to a lower risk of graft failure (−2.9%) and iatrogenic acute glaucoma (−12.0%) in the iOCT-optimized protocol. However, the analysis shows a high uncertainty regarding effect sizes for all adverse events and non-inferiority cannot be assessed for these stratified outcomes, because the study was not powered on these separate adverse events.

No significant differences were found between the control group and the iOCT group regarding secondary clinical outcomes at 3 and 6 months postoperative (table 2). In particular, the ECD loss, RFNL, and postoperative IOP did not differ between both groups and harmful long-term effects of prolonged overpressure thus appear unlikely in patients without prior retinal nerve damage.

### Usefulness of intra-operative OCT for surgical decision-making and surgical time

In 35 surgeries the iOCT was utilized, including 2 cross-over cases in an attempt to save the grafts. The graft orientation in these cases was particularly difficult to assess. The use of iOCT salvaged the graft in one case. The other graft was correctly positioned though eventually developed a graft failure, presumably because of repeated manipulation. None of the iOCT-group cases exhibited interface irregularities or graft detachment at the end of surgery. The obligatory verification scan in the control group revealed peripheral detachment of the graft in one case, resulting in repositioning of the graft and subsequent over-pressure for another 4 minutes. Notwithstanding, this case developed a detachment for which a rebubbling was performed.

Surgeons reported that the iOCT benefited decision-making in 14 of 35 cases (40%); in all cases (14/14) iOCT aided determining graft orientation (incl. 8 grafts inserted upside-down) and in 21% (3/14) iOCT aided unfolding and positioning of the graft. The median time the iOCT was used measured 2 minutes and 52 seconds (IQR: 03:43, range: 00:19 – 23:40). In 28 cases the image quality was considered good (85%), in four cases acceptable (12%), and in one case poor (3%). The graft unfolding grade was significantly associated with surgeon reported iOCT-aided surgical decision-making (Supplementary table 2, P=0.011, Fisher exact test); in cases with a complicated graft unfolding the iOCT proved to benefit surgical decision-making. Notwithstanding, graft unfolding grade did not differ between both treatment arm (Supplementary table 2, P=0.474, Fisher exact test).

As expected, refraining from prolonged overpressure resulted in a shorter mean surgical skin-to-skin time in the iOCT group compared to the control group (mean difference: 4.90 minutes, SD±2.51, -13%). In addition, the mean graft unfolding time in the iOCT group was 1.68 minutes shorter (SD±0.85, - 26.8%). An overview of the duration of the main surgical steps is shown in table 4.

**Table 4.** Overview of surgical times Manually scored and video-graded by two independent observers

|  | Conventional treatment (control group) | iOCT-optimized treatment | Relative difference (%) | 95% CI |
| --- | --- | --- | --- | --- |
| Surgical skin-to-skin time (minutes), mean (SD) | 37.62 (10.09) | 32.72 (10.99) | -13.0 | -0.36 – 10.18 |
| Overpressure time(minutes), mean (SD) | 9.73 (1.94) | 2.73 (1.29) | -71.9 | 6.19 – 7.82 |
| unfolding time, minutes, (minutes), mean (SD) | 6.26 (8.13) | 4.58 (5.35) | -26.8 | -1.75 – 5.10 |
| Graft preparation time<sup>1</sup>, (minutes), mean (SD) | 6.83 (1.95) | 6.97 (2.01) | 2.0 | -1.25 – 0.98 |
| Descemetorhexis time, (minutes), mean (SD) | 4.35 (4.41) | 5.90 (4.84) | 35.6 | -3.88 – 0.78 |

SD: standard deviation, iOCT; intraoperative optical coherence tomography
<sup>1</sup> Grafts preparation time for surgeries in Leuven was not available.

## Discussion

In this study we found that an iOCT-optimized DMEK surgical protocol with iOCT-guidance and refraining from over pressurizing was non-inferior compared to a conventional protocol, with no iOCT-guidance and standard 8 minutes of over pressure. Our results do not support the perceived benefit of overpressure to promote graft adherence. Though the independent effect of iOCT use on surgical safety could not be reliably estimated, the benefits of our iOCT-optimized protocol are a shorter surgical skin-to-skin time (−13%) and assisted surgical decision making (40% of cases). Furthermore, the access to iOCT and its improved visualization proved crucial during surgery in 9% of cases in the control group (2 crossovers and 1 validation scan with observed intra-operative graft detachment).

The causes of graft detachments are considered multifactorial and a large body of research reported on risk factors, such as donor and recipient characteristics, and intraoperative factors such as overpressure of the globe.^10,31–33^ Over-pressuring during surgery is considered by some as a protective factor against graft detachments^15,21^, whereas two cohort studies did not support this.^19,20^ Our study is the first head-to-head comparison of over-pressuring in DMEK surgery, and whilst graft detachments were prevalent in both treatment arms, our data do not support the notion that over-pressure prevents graft detachments nor rebubbling procedures. Apparently, the incidence of detachments is driven by other factors than assessed in this clinical trial, such as patient compliance with given instructions on immobilization, or different anterior chamber tamponade strategies (e.g. long-term complete air-fill).^34,35^ Both could be interesting entry points for follow-up clinical studies.

Overall, the prevalence of adverse events did not differ significantly or materially between both treatment arms (iOCT-group, n=13; control group, n=13). However, the nature of the separate adverse events differed. For instance, the incidence of iatrogenic glaucoma was higher in the prolonged pressurization group. Potentially, a prolonged high pressure in the AC during surgery forces small amounts of gas behind the iris, subsequently leading to an episode of post-operative acute glaucoma, though the exact physiologic process remains unclear. Refraining from overpressure may help to reduce the incidence of postoperative iatrogenic acute glaucoma and benefit patients with pre-existing glaucoma. A harmful effect of prolonged overpressure in our non-glaucomatous population was not identified, though this is an interesting question for follow-up clinical studies. Interestingly, graft detachments occurred at an equal rate (n=17 vs. n=16) and the areas of detachment were of comparable size, though rebubbling procedures were performed more often in the iOCT group. The cause of this difference remain unclear as our study was not designed to assess nor explore predictors for clinical decision making regarding rebubbling procedures. The decision to re-adhere a graft is made by the surgeon which may be related to contextual factors not assessed in this clinical trial, such as location of detachment, tissue- or patient characteristics.

The use of iOCT benefitted the surgical decision-making process in 40% of cases. This finding is consistent with results from comparable studies, including our pilot study and the landmark PIONEER and DISCOVER studies.^3–6,24,25^ Similar to these studies our surgeons reported that the iOCT imaging was particularly advantageous for assessing graft orientation and in lesser degree during the unfolding of the graft. Interestingly, we found a significant association between reported iOCT-aided surgical decision-making and the graded unfolding difficulty. This makes sense, as the circumstances and causes which make graft orientation difficult to assess (e.g., poor visualization, graft geometry and tissue properties) may also increase the difficulty of unfolding the graft^28^ Hallahan et al. proposed that the iOCT-image may influence the aggressiveness of manipulations.^36^ Our data indicate that the iOCT is more utilized and perceived more useful in difficult cases, though not directly related to graft unfolding difficulty nor aggressiveness of manipulations, since these occurred equally in both treatment arms.

The surgical skin-to-skin time was 13% shorter in the iOCT group, which was expected due to refraining from overpressure in the iOCT-optimized protocol. In addition, in line with similar reports we found that the iOCT enables the surgeon in a 26% faster unfolding and positioning of the graft. Though not assessed in this study a shorter duration of unfolding and positioning the graft may be related to less manipulation of the graft and improved graft viability and survival.^3–5^ Efficiency gains from refraining from overpressure and a faster unfolding time may be offset by the surgeon taking time to assess the iOCT images. We recorded the time iOCT was switched on (median 2:52, IQR 3:43, range 00:19 – 23:40), though the actual time spent by the surgeon assessing iOCT images is difficult to measure. Evidently, this assessment time is much shorter than the total iOCT time. Future development in automated image analysis may aid to reduce this offset.^1,37^

Long-term follow-up results appeared comparable for both groups. Endothelial cell density is a major determinant for long-term graft survival. The postoperative ECD loss was slightly lower in the conventional protocol compared with the iOCT-optimized protocol in this study, albeit not statistically significant. In addition to other reports, the combined results may suggest that ECD is not affected by prolonged pressuring of the globe and thus not related to reduced long-term graft viability.^21,23^ In our study no sequelae of prolonged pressurization were found at follow-up in regard to postoperative IOP or retinal nerve fiber layer damage, which is in line with the report from Fortune et al.^38^

The relative costs of the iOCT-system warrant discussion given the non-inferiority of our iOCT-optimized protocol. We aimed to quantify the benefit of an iOCT guided surgical protocol, which shortened surgical times considerably. iOCT improved surgical decision making, proved indispensable in selected cases according to the surgeons, and enabled adeqate management of intrasurgical events. However, this is not reflected in overall post-operative adverse event rates and clinical outcomes. Although one could hypothesize that refraining from overpressure is also non-inferior regardless intraoperative imaging, this should first be confirmed by clinical studies. Applications that benefit the surgical process beyond DMEK surgery are taking shape, which could be considered a justification of investing in iOCT technology.

Several limitations should be addressed. The study design evaluated two important outcomes in DMEK surgery: the use of iOCT and the use of overpressure. The partial effect of these individual factors is difficult to estimate reliably due to the introduced multi-collinearity. In selected cases, iOCT proved indispensable for the surgeon to complete the surgery successfully, and many reports highlight this benefit of iOCT, though it is not feasible to power a trial on these rare cases and outcomes.^1^ Additionally, when iOCT is available at a center, withholding this technology from complex cases (e.g. clouded corneas) is considered unethical.^39^ We firmly believe that new innovations should be tested on endpoints relevant for patients, and assessing process-related outcomes (e.g. surgical time) can only be secondary to a primary outcome that relates directly to the patient (e.g. surgical safety). In addition, we attempted to assess the actual IOP during over-pressurization using a hand-held (rebound) tonometer. These measurements proved extremely variable (data not shown) and not related to other clinical signs of over-pressure (firm globe, pupil dilation). Animal studies with custom-made intra-ocular manometers exist^40^, but we assume that the measurement of IOP in air/gas filled eyes with conventional certified devices is not feasible. Another consideration is in the interpretation of outcomes regarding graft detachments and rebubbling events. In our study protocol, we listed rebubbling as a primary outcome due to its relevance from a patient perspective, though advancing insights let to the conclusion that a graft detachment is a more objective and quantifiable outcome. We therefore reported both and acknowledge that the decision to re-adhere a graft is made by the surgeon. In the study protocol we did not prescribe strict guidelines on when to intervene with a graft detachment to preserve clinical discretion, which could also be considered a limitation.

In conclusion, iOCT-guided DMEK surgery refraining from prolonged over-pressurizing was proven non-inferior to a conventional approach, though it did not reduce the overall rate of post-operative adverse events. Surgery times were reduced overall by 13% and the iOCT resulted in a 27% reduction of unfolding time. Surgeons reported a benefit of iOCT in 40% of cases. Follow-up studies should elucidate the multi-factorial origin of graft detachment after lamellar corneal transplant surgery.

## Data Availability

All data produced in the present study are available upon reasonable request to the authors

## Acknowledgements

The authors would like to thank Suryan Dunker, MD, and Ingeborg de Vries, BSc, for their assistance with the trial measurements, Peter Zuithoff, PhD, of the Julius Center for Health Sciences, for assistance and validation of the statistical analysis, Hanad Jimale, BSc, for his assistance in grading the surgical videos, and Antoon van den Bogaerdt, PhD, from ETB-Bislife for his aid in providing donor tissue to all participating centers.

## References

1. Muijzer MB, Schellekens Pawj, Beckers HJM, de Boer JH, Imhof SM, Wisse RPL. Clinical applications for intraoperative optical coherence tomography: a systematic review. Eye. 2021;(June):1–13. doi:10.1038/s41433-021-01686-9

2. Ehlers JP, Modi YS, Pecen PE, et al. The DISCOVER Study 3-Year Results: Feasibility and Usefulness of Microscope-Integrated Intraoperative OCT during Ophthalmic Surgery. Ophthalmology. 2018;125(7):1014–1027. doi:10.1016/j.ophtha.2017.12.037

3. Ehlers JP, Dupps WJ, Kaiser PK, et al. The Prospective Intraoperative and Perioperative Ophthalmic ImagiNg with Optical CoherEncE TomogRaphy (PIONEER) Study: 2-year results. Am J Ophthalmol. 2014;158(5):999–1007. doi:10.1016/j.ajo.2014.07.034

4. Posarelli C, Sartini F, Casini G, et al. What Is the Impact of Intraoperative Microscope-Integrated OCT in Ophthalmic Surgery? Relevant Applications and Outcomes. A Systematic Review. J Clin Med. 2020;9(6). doi:10.3390/jcm9061682

5. Saad A, Guilbert E, Grise-Dulac A, Sabatier P, Gatinel D. Intraoperative OCT-Assisted DMEK: 14 Consecutive Cases. Cornea. 2015;34(7):802–807. doi:10.1097/ICO.0000000000000462

6. Patel AS, Goshe JM, Srivastava SK, Ehlers JP. Intraoperative Optical Coherence Tomography– Assisted Descemet Membrane Endothelial Keratoplasty in the DISCOVER Study: First 100 Cases. Am J Ophthalmol. 2020;210(February 2018):167–173. doi:10.1016/j.ajo.2019.09.018

7. Cost B, Goshe JM, Srivastava S, Ehlers JP. Intraoperative optical coherence tomography-assisted descemet membrane endothelial keratoplasty in the DISCOVER study. Am J Ophthalmol. 2015;160(3):430–437. doi:10.1016/j.ajo.2015.05.020

8. Muijzer MB, Soeters N, Godefrooij DA, van Luijk CM, Wisse RPL. Intraoperative Optical Coherence Tomography-Assisted Descemet Membrane Endothelial Keratoplasty: Toward More Efficient, Safer Surgery. Cornea. 2020;39(6):674–679. doi:10.1097/ICO.0000000000002301

9. Dunker SL, Dickman MM, Wisse RPL, et al. Descemet Membrane Endothelial Keratoplasty versus Ultrathin Descemet Stripping Automated Endothelial Keratoplasty: A Multicenter Randomized Controlled Clinical Trial. Ophthalmology. 2020;127(9):1152–1159. doi:10.1016/j.ophtha.2020.02.029

10. Dunker SL, Veldman MHJ, Winkens B, et al. Real-World Outcomes of DMEK: A Prospective Dutch registry study. Am J Ophthalmol. 2021;222:218-225. doi:10.1016/j.ajo.2020.06.023

11. Stuart AJ, Romano V, Virgili G, Shortt AJ. Descemet’s membrane endothelial keratoplasty (DMEK) versus Descemet’s stripping automated endothelial keratoplasty (DSAEK) for corneal endothelial failure. Cochrane Database Syst Rev. 2018;2018(6). doi:10.1002/14651858.CD012097.pub2

12. Dunker S, Winkens B, Van Den Biggelaar F, Nuijts R, Kruit PJ, Dickman M. Rebubbling and graft failure in Descemet membrane endothelial keratoplasty: a prospective Dutch registry study. Br J Ophthalmol. 2021;(October 2011):1-7. doi:10.1136/bjophthalmol-2020-317041

13. Singh A, Zarei-Ghanavati M, Avadhanam V, Liu C. Systematic Review and Meta-Analysis of Clinical Outcomes of Descemet Membrane Endothelial Keratoplasty Versus Descemet Stripping Endothelial Keratoplasty/Descemet Stripping Automated Endothelial Keratoplasty. Cornea. 2017;36(11):1437–1443. doi:10.1097/ICO.0000000000001320

14. Li S, Liu L, Wang W, et al. Efficacy and safety of Descemet’s membrane endothelial keratoplasty versus Descemet’s stripping endothelial keratoplasty: A systematic review and meta-analysis. PLoS One. 2017;12(12):1–21. doi:10.1371/journal.pone.0182275

15. Terry MA, Aldave AJ, Szczotka-Flynn LB, et al. Donor, Recipient, and Operative Factors Associated with Graft Success in the Cornea Preservation Time Study. Ophthalmology. 2018;125(11):1700–1709. doi:10.1016/j.ophtha.2018.08.002

16. Dirisamer M, Van Dijk K, Dapena I, et al. Prevention and management of graft detachment in descemet membrane endothelial keratoplasty. Arch Ophthalmol. 2012;130(3):280–291. doi:10.1001/archophthalmol.2011.343

17. Terry MA, Straiko MD, Veldman PB, et al. Standardized DMEK Technique: Reducing Complications Using Prestripped Tissue, Novel Glass Injector, and Sulfur Hexafluoride (SF6) Gas. Cornea. 2015;34(8):845–852. doi:10.1097/ICO.0000000000000479

18. Santander-García D, Peraza-Nieves J, Müller TM, et al. Influence of Intraoperative Air Tamponade Time on Graft Adherence in Descemet Membrane Endothelial Keratoplasty. Cornea. 2019;38(2):166–172. doi:10.1097/ICO.0000000000001795

19. Price FW, Price MO. Descemet’s stripping with endothelial keratoplasty in 200 eyes. Early challenges and techniques to enhance donor adherence. J Cataract Refract Surg. 2006;32(3):411–418. doi:10.1016/j.jcrs.2005.12.078

20. Heinzelmann S, Böhringer D, Haverkamp C, et al. Influence of postoperative intraocular pressure on graft detachment after descemet membrane endothelial keratoplasty. Cornea. 2018;37(11):1347–1350. doi:10.1097/ICO.0000000000001677

21. Pilger D, Wilkemeyer I, Schroeter J, Maier AKB, Torun N. Rebubbling in Descemet Membrane Endothelial Keratoplasty: Influence of Pressure and Duration of the Intracameral Air Tamponade. Am J Ophthalmol. 2017;178:122–128. doi:10.1016/j.ajo.2017.03.021

22. Schmeckenbächer N, Frings A, Kruse FE, Tourtas TL. Role of Initial Intraocular Pressure in Graft Adhesion After Descemet Membrane Endothelial Keratoplasty. 2016;0(0):1–4.

23. Fajgenbaum MAP, Hollick EJ. Does Same-Day Postoperative Increased Intraocular Pressure Affect Endothelial Cell Density After Descemet Membrane Endothelial Keratoplasty? Cornea. 2018;37(12):1484–1489. doi:10.1097/ICO.0000000000001762

24. Janson BJ, Alward WL, Kwon YH, et al. Glaucoma-associated corneal endothelial cell damage: A review. Surv Ophthalmol. 2018;63(4):500–506. doi:10.1016/j.survophthal.2017.11.002

25. Khng C, Packer M, Fine IH, Hoffman RS, Moreira FB. Intraocular pressure during phacoemulsification. J Cataract Refract Surg. 2006;32(2):301–308. doi:10.1016/j.jcrs.2005.08.062

26. Schulz KF, Altman DG, Moher D. CONSORT 2010 Statement: updated guidelines for reporting parallel group randomised trials. BMJ. 2010;340. doi:10.1136/bmj.c332

27. Dapena I, Moutsouris K, Droutsas K, Ham L, van Dijk K, Melles G. Standardized “No-Touch” Technique for Descemet Membrane Endothelial Keratoplasty. Arch Ophthalmol. 2011;129(1):88–94. doi:10.1001/archophthalmol.2010.334

28. Maier AKB, Gundlach E, Schroeter J, et al. Influence of the difficulty of graft unfolding and attachment on the outcome in descemet membrane endothelial keratoplasty. Graefe’s Arch Clin Exp Ophthalmol. 2015;253(6):895–900. doi:10.1007/s00417-015-2939-9

29. Beck RW, Moke PS, Turpin AH, et al. A computerized method of visual acuity testing: Adaptation of the early treatment of diabetic retinopathy study testing protocol. Am J Ophthalmol. 2003;135(2):194–205. doi:10.1016/S0002-9394(02)01825-1

30. Austin PC. Absolute risk reductions, relative risks, relative risk reductions, and numbers needed to treat can be obtained from a logistic regression model. J Clin Epidemiol. 2010;63(1):2–6. doi:10.1016/j.jclinepi.2008.11.004

31. Parekh M, Leon P, Ruzza A, et al. Graft detachment and rebubbling rate in Descemet membrane endothelial keratoplasty. Surv Ophthalmol. 2018;63(2):245–250. doi:10.1016/j.survophthal.2017.07.003

32. Weller JM, Schlötzer-Schrehardt U, Tourtas T, Kruse FE. Influence of Ultrastructural Corneal Graft Abnormalities on the Outcome of Descemet Membrane Endothelial Keratoplasty. Am J Ophthalmol. 2016;169:58–67. doi:10.1016/j.ajo.2016.06.013

33. Quilendrino R, Rodriguez-Calvo de Mora M, Baydoun L, et al. Prevention and Management of Graft Detachment in Descemet Membrane Endothelial Keratoplasty. Arch Ophthalmol. 2012;130(3):280–291. doi:10.1001/archophthalmol.2011.343

34. Gonzalez A, Price FW, Price MO, Feng MT. Prevention and Management of Pupil Block after Descemet Membrane Endothelial Keratoplasty. Cornea. 2016;35(11):1391–1395. doi:10.1097/ICO.0000000000001015

35. Cirkovic A, Beck C, Weller JM, Kruse FE, Tourtas T. Anterior chamber air bubble to achieve graft attachment after DMEK: Is bigger always better? Cornea. 2016;35(4):482–485. doi:10.1097/ICO.0000000000000753

36. Hallahan KM, Cost B, Goshe JM, Dupps WJ, Srivastava SK, Ehlers JP. Intraoperative Interface Fluid Dynamics and Clinical Outcomes for Intraoperative Optical Coherence Tomography– Assisted Descemet Stripping Automated Endothelial Keratoplasty From the PIONEER Study. Am J Ophthalmol. 2017;173:16–22. doi:10.1016/j.ajo.2016.09.028

37. Xu D, Dupps WJJ, Srivastava SK, Ehlers JP. Automated volumetric analysis of interface fluid in descemet stripping automated endothelial keratoplasty using intraoperative optical coherence tomography. Invest Ophthalmol Vis Sci. 2014;55(9):5610–5615. doi:10.1167/iovs.14-14346

38. Fortune B, Yang H, Strouthidis NG, et al. The effect of acute intraocular pressure elevation on peripapillary retinal thickness, retinal nerve fiber layer thickness, and retardance. Investig Ophthalmol Vis Sci. 2009;50(10):4719–4726. doi:10.1167/iovs.08-3289

39. Muijzer MB, Kroes HY, Van Hasselt P, Wisse RPL. Bilateral posterior lamellar corneal transplant surgery in an infant of 17 weeks old: surgical challenges and the added value of intraoperative optical coherence tomography. Authorea. Published online 2021.

40. Cheng W, Liu L, Yu S, et al. Real-Time Intraocular Pressure Measurements in the Vitreous Chamber of Rabbit Eyes During Small Incision Lenticule Extraction (SMILE). Curr Eye Res. 2018;43(10):1260–1266. doi:10.1080/02713683.2018.1485949

41. Wisse RPL, Muijzer MB, Hoven CMW, Van Luijk CM, Vink G, Frank LE. A machine learning approach to identify predictors of adverse events in posterior lamellar keratoplasty; a nationwide registry study. Acta Ophthalmol. 2021;99(SUPPL 266):6. doi:10.1111/aos.

