## Supplementary table 1 for "Outcomes of the Advanced Visualization In Corneal Surgery Evaluation (ADVISE) trial; a non-inferiority randomized control trial to evaluate the use of intraoperative OCT during Descemet membrane endothelial keratoplasty"

Supplementary table 1. Logistic regression models for incidence of total adverse event rate

| | Unadjusted model | | Adjusted model | |
|---|---|---|---|---|
| **Variables** | **OR (95%CI)** | **P** | **OR (95%CI)** | **P** |
| iOCT-optimized protocol | 1.036 (0.454 - 2.366) | 0.933 | 0.976 (0.421 – 2.263) | 0.955 |
| Study site 2[1] | - | - | 0.123 (0.016 – 0.953) | 0.045 |
| Study site 3[1] | - | - | 0.634 (0.221 – 1.819) | 0.396 |

1 Reference: study site 1

iOCT: intraoperative optical coherence tomography
