## Supplementary table 2 for "Outcomes of the Advanced Visualization In Corneal Surgery Evaluation (ADVISE) trial; a non-inferiority randomized control trial to evaluate the use of intraoperative OCT during Descemet membrane endothelial keratoplasty"

Supplementary table 2. Overview of graft unfolding grade in both treatment arms

| Treatment arm / Graft unfolding grade[1] | | I | II | III | IV | P-value[2] |
|---|---|---|---|---|---|---|
| Conventional protocol, **n** | | 4 | 14 | 4 | 11 | 0.474 |
| iOCT-optimized protocol, **n** | | 7 | 14 | 1 | 9 | |
| | iOCT aided surgical decision-making | 1 | 4 | 1 | 7 | 0.011 |
| | iOCT did not aid surgical decision-making | 6 | 10 | 0 | 2 | |

[1] Graft unfolding grade is classified in 4 grades depending on the required manipulation and time to unfold/position the graft. Grade I: graft lamella primarily oriented correctly in the anterior chamber, straight and direct unfolding and centering; Grade II: slightly complicated, indirect unfolding and centering (duration less than five min); Grade III: difficult indirect unfolding and centering (duration longer than five min), repeated air injection with BSS exchange necessary; Grade IV: direct manipulation of the graft lamella for unfolding and centering by cannula or forceps.

[2] Fisher exact test
